# Association between sleep and seroconversion after vaccination with inactivated SARS-CoV-2 in pregnant women

**DOI:** 10.1101/2023.06.04.23290946

**Authors:** Xue Han, Wenhan Yang, Xi Zhang, Baolan Chen, Xi Fu, Jitian Huang, Yingxia Xu, Yajie Gong, Qingsong Chen

## Abstract

To assess the association between sleep and seroconversion after receipt of two doses of inactivated severe acute respiratory syndrome-coronavirus-2 (SARS-CoV-2) vaccines in pregnant women. The serum level of immunoglobulin (Ig)G antibodies against the nucleic acids of SARS-CoV-2 was measured. Logistic regression was used to analyze the association between sleep and seroconversion. After two doses of SARS-CoV-2 vaccine, 41.2% of the study cohort reached seroconversion. Analysis revealed that pregnant women with poor quality of sleep had a lower serum level of IgG antibodies (*P* = 0.008, 95%CI = 0.285–0.826) and that sleeping late at night (SLaN) may be a risk factor for a low serum level of IgG antibodies (*P* = 0.025, 95%CI = 0.436–0.946). Besides sleep, age and the time since vaccination were important influences on seroconversion. A stratified analysis revealed that the effects of sleep quality and SLaN on seroconversion occurred mainly in pregnant women aged <35 years. Thus, sleep quality and SLaN can affect the serum level of IgG antibodies in pregnant women after vaccination with inactivated SARS-CoV-2.

## Introduction

The coronavirus disease-2019 (COVID-19) pandemic was once an important public-health issue worldwide despite aggressive government responses. Mass vaccination is a key measure to control the spread of this outbreak. Dozens of severe acute respiratory syndrome-coronavirus disease-2 (SARS-CoV-2) vaccines have been approved for use, with >1.2 billion doses administered worldwide as of August 2022^[1]^. Countries used to adopt home-quarantine methods to control the spread of COVID-19, in addition to large-scale vaccination.

The symptoms of COVID-19 infection and the side effects of vaccination may lead to sleep problems, but whether it is also related to other factors of the vaccine is unknown. Sleep is a state of total rest and active neuromodulation of the central nervous system required by the body to eliminate fatigue. Several studies have shown that sleep can be involved (directly or indirectly) in regulation of the immune system through its effects on the nervous system and neuroendocrine system^[2,3]^. Maintenance of many immune functions is dependent on circadian rhythms and regular sleep. Sleep can affect various components of the immune system, and sleep disturbance can cause changes in the number and function of immune cells.

Clinical findings have shown that sleep deprivation affects expression of proinflammatory factors in central and peripheral systems in humans. Five nights of sleep restriction has been shown to increase circulating levels of the proinflammatory factors interleukin (IL)-1β, IL-6, and IL-17, which remain at high levels even after 2 nights of sleep recovery^[4]^. In a prospective cohort study from Greece, in which all enrolled participants received two doses of a vaccine based on mRNA SARS-CoV-2, insomnia was found to be associated with a lower serum level of immunoglobulin (Ig)G antibodies, but that study did not involve pregnant women^[5].^

Physiological changes during pregnancy have significant effects on the immune system and respiratory system. The effect on the immune system after vaccination is influenced by various factors. However, few studies have investigated the effect of sleep on the immune system of pregnant women after administration of a vaccine based on inactivated SARS-CoV-2.

Here, we investigated the effect of sleep on the serum level of IgG antibodies in pregnant women after receipt of two doses of inactivated SARS-CoV-2 vaccine.

## Methods

### Study design

We collected information from the electronic medical records of pregnant women who gave birth in three hospitals in Guangzhou from December 2021 to March 2022. These women were given questionnaires to complete. All participants provided written informed consent. The study protocol complied with the Declaration of Helsinki 1964 and its later amendments. The data they provided included demographic information as well as information on basic behavioral lifestyle, including marital status, educational level, occupation, age (stratified by the advanced maternal age defined by the World Health Organization), body mass index, time of last menstruation, sleep status (duration of nighttime sleep, “napping”, sleeping late at night (defined as go to bed past 11 p.m, SLaN), sleep disturbance, sleep quality), tobacco smoking, alcohol consumption, pregnancy, disease history, and the time since vaccination (defined as the time interval between the last dose of vaccine and blood collection).

To exclude the effect of different doses of vaccine on the serum level of IgG antibodies, only pregnant women who received two doses of inactivated SARS-CoV-2 vaccine were included in this study. Pregnant women who had been infected or were infected with SARS-CoV-2 after enrollment and who received non-inactivated vaccine were excluded.

### Collection and analyses of samples

After enrollment, 5 mL of venous blood was collected, centrifuged to separate the serum, and stored at −80°C. IgG antibodies against the nucleic acids of SARS-CoV-2 were detected using a chemiluminescence kit (Bioscience, Chongqing, China). A serum level of IgG antibodies <1.00 signal/cutoff (S/CO) ratio was considered negative. A serum level of IgG antibodies ≥1.00 S/CO was considered positive.

### Statistical analyses

Descriptive analysis was used to denote demographic features. Factors that may affect the serum level of IgG antibodies were identified using the chi-squared test or rank-sum test. Logistic regression analysis was employed to analyze the relationship between sleep-related factors and antibody positivity. Statistical analyses were undertaken using SPSS 25.0 (IBM, Armonk, NY, USA). P < 0.05 (two-sided) was considered significant.

## Results

### Demographic and clinical characteristics

The study cohort comprised 650 pregnant women aged 18–43 years who had received two doses of inactivated SARS-CoV-2 vaccine before or during pregnancy. The number of pregnant women who had two doses of SARS-CoV-2 vaccine before and during pregnancy was 544 (83.7%) and 106 (16.3%), respectively.

A total of 440 (67.6%) pregnant women had suffered from a sleep disturbance in the previous year. These sleep disturbance included: difficulty in falling asleep (unable to fall asleep within 30 min); awakening readily or early at night; frequent visits to the toilet at night; nightmares; excessive dreaming. Seventy-nine pregnant women were aged >35 years (12.1%) (**Table 1**).

**Table 1.**
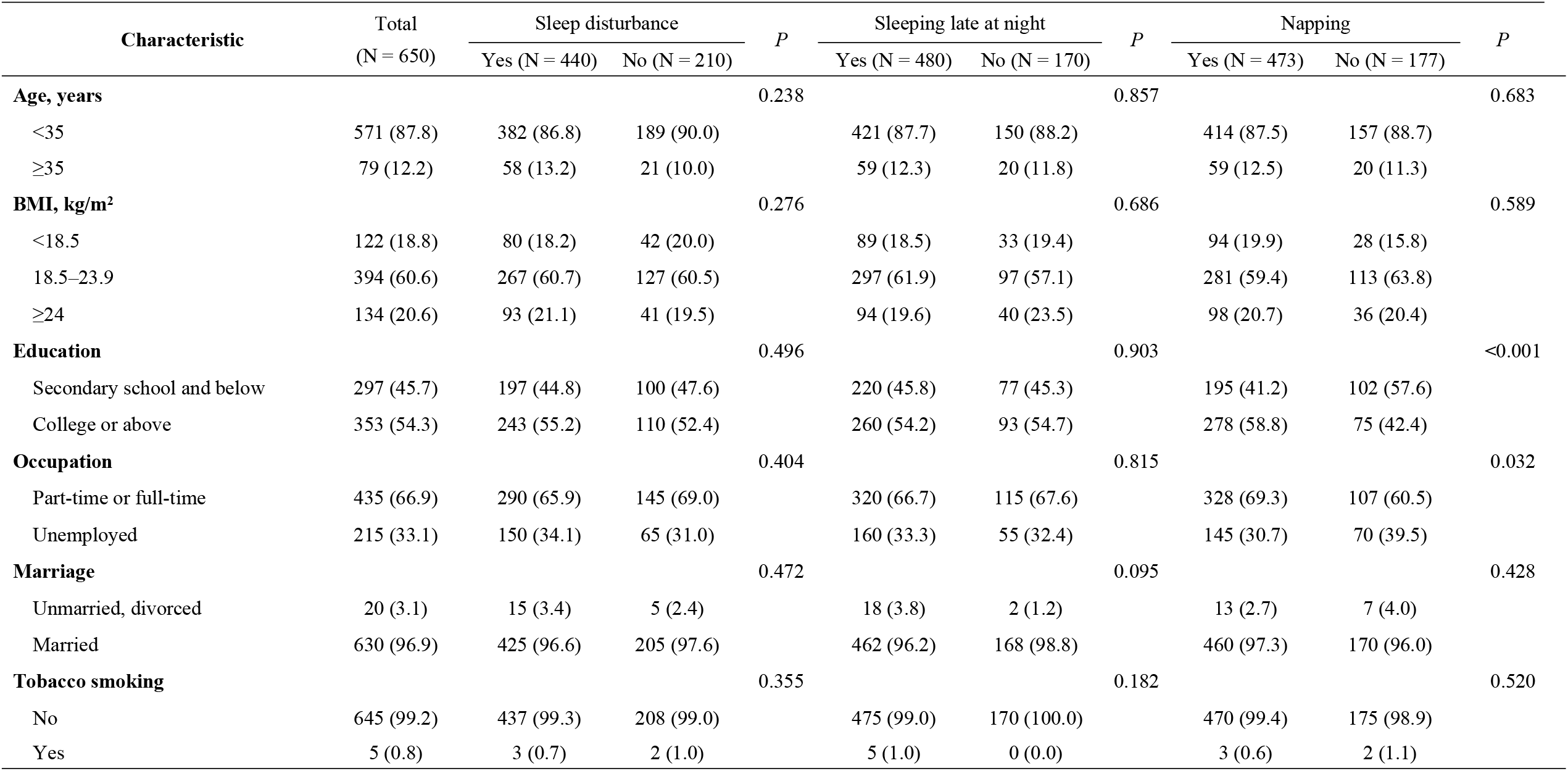
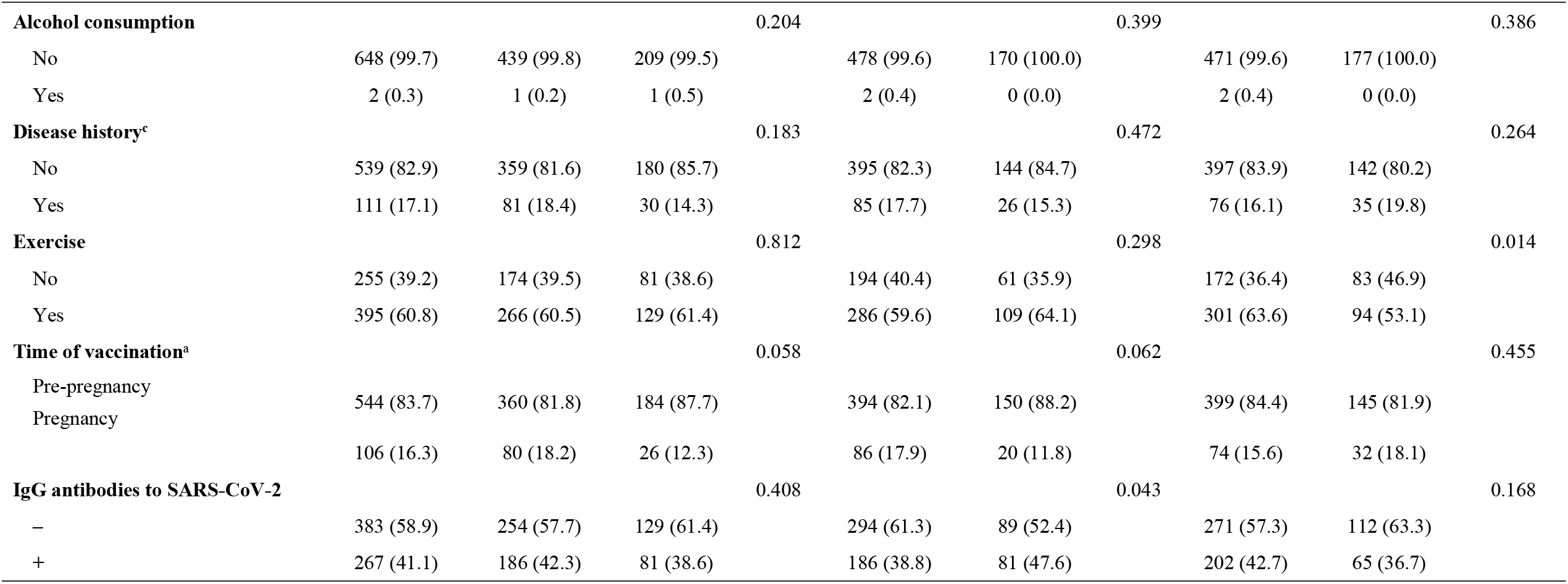
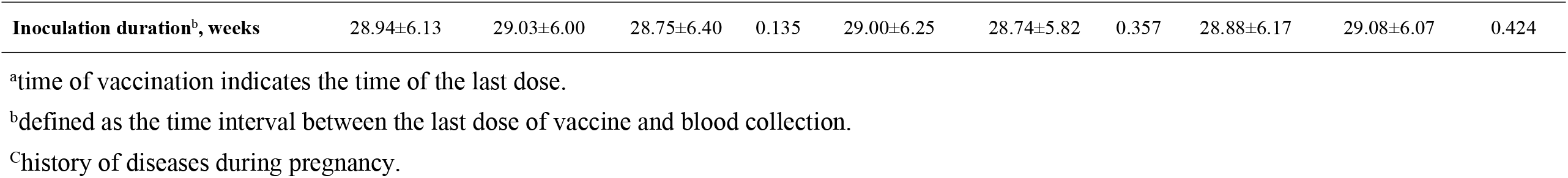
Demographic characteristics of participants.

### Relationship between the serum concentration of IgG antibodies and sleep

“Seroconversion” is the development of specific antibodies in the blood serum as a result of infection or immunization, including vaccination. We discovered that, after vaccination, 41.2% of pregnant women had produced IgG antibodies against the nucleic acids of SARS-CoV-2. Most of the enrolled pregnant women had the habit of SLaN (73.8%) and taking a nap (72.8%). The duration of nighttime sleep was 8.51±1.14 h and the duration of a single nap was 0.81±0.81 h. Univariate analysis revealed a significant difference between education level, occupation, and exercise status and napping (*P* < 0.05). A significant difference between the prevalence of SLaN and production of IgG antibodies was documented (*P* = 0.043)(**Table 1**).

Logistic regression analysis revealed a significant difference between sleep quality, SLaN, and production of IgG antibodies (*P* < 0.05)(**Table 2**). We discovered that the prevalence of seroconversion was lower in pregnant women who had poor quality of sleep (*P* = 0.008, 95%CI 0.285−0.826) and that SLaN may be a risk factor for low production of IgG antibodies in pregnant women (*P* = 0.025, 95%CI 0.436−0.946), which persisted even after adjustment for confounders. In addition to the significant difference between sleep factors and production of IgG antibodies, age (*P* = 0.016, 95%CI 0.307−0.885) and the time since vaccination (defined as the time interval between the last dose of vaccine and blood collection) (*P*< 0.001, 95%CI 0.912−0.967) were associated with seroconversion.

**Table 2.** Multifactorial logistic regression analysis of antibody positivity.

| Characteristic | IgG antibodies to SARS-CoV-2 |  |
| --- | --- | --- |
|  | OR (95%CI)<sup>a</sup> | *P* |
| **Age group, years** |  |  |
| <35 | 1.00 |  |
| ≥35 | 0.522 (0.307–0.885) | **0.016** |
| **Inoculation duration, weeks** | 0.939 (0.912–0.967) | **<0.001** |
| **Sleep quality** |  |  |
| Good | 1.00 |  |
| Poor | 0.485 (0.285–0.826) | **0.008** |
| **Duration of nighttime sleep, h** |  |  |
| <8 | 1.00 |  |
| ≥8 | 0.774 (0.535–1.119) | 0.173 |
| **Sleep disturbance** | 1.332 (0.932–1.903) | 0.116 |
| **Napping** | 1.257 (0.862–1.834) | 0.234 |
| **Sleeping late at night** | 0.642 (0.436–0.946) | **0.025** |

OR, odds ratio; 95% CI, 95% confidence interval.
<sup>a</sup>Adjusted for BMI, education, occupation, tobacco smoking, alcohol consumption, exercise, time of vaccination, history of diseases during pregnancy.

For the further exploring the differences between SLaN, sleep quality, and the serum level of IgG antibodies in pregnant women of different age groups and at the time since vaccination, a stratified analysis was conducted within two groups: pregnant women aged 35 years (**Table 3**); 24 weeks after vaccination (**Table 4**). The effects of sleep quality and SLaN on the serum level of IgG antibodies in pregnant women were found mainly in the group younger than 35 years. No relationship between sleep quality, SLaN, and the serum level of IgG antibodies was found in the group vaccinated <24 weeks previously. However, in the group vaccinated >24 weeks previously, there was an association between SLaN and the serum level of IgG antibodies, but not significantly so (*P* = 0.053).

**Table 3.**
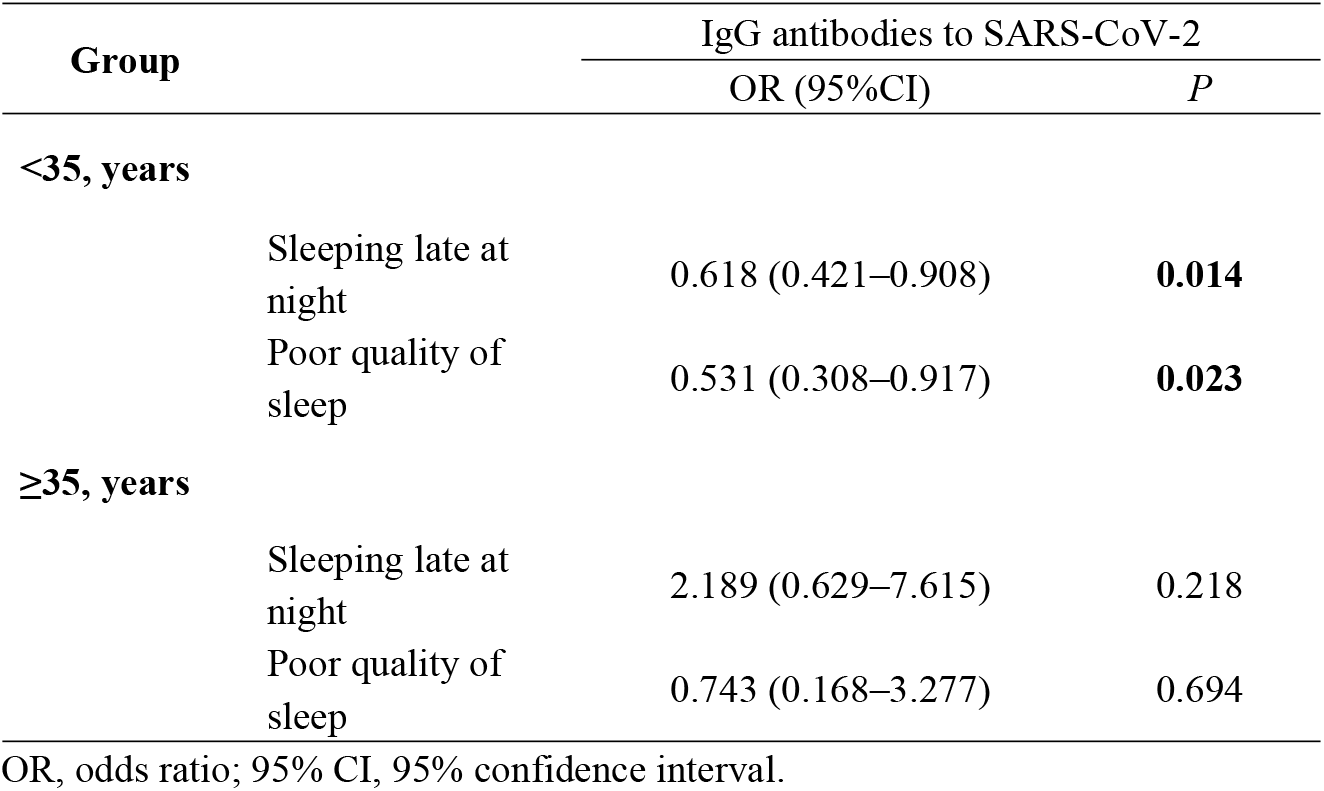
Association between pregnancy, sleep, and antibody level by age.

| Group | IgG antibodies to SARS-CoV-2 |  |
| --- | --- | --- |
|  | OR (95%CI) | *P* |
| **<35, years** |  |  |
| Sleeping late at night | 0.618 (0.421–0.908) | **0.014** |
| Poor quality of sleep | 0.531 (0.308–0.917) | **0.023** |
| **≥35, years** |  |  |
| Sleeping late at night | 2.189 (0.629–7.615) | 0.218 |
| Poor quality of sleep | 0.743 (0.168–3.277) | 0.694 |

OR, odds ratio; 95% CI, 95% confidence interval.

**Table 4.** Association between pregnancy, sleep, and antibody level according to the time since vaccination.

| Group | IgG antibodies to SARS-CoV-2 |  |
| --- | --- | --- |
|  | OR (95%CI) | *P* |
| <24 weeks |  |  |
| Sleeping late at night | 0.758 (0.307–1.872) | 0.548 |
| Poor sleep quality | 0.412 (0.148–1.146) | **0.089** |
| ≥24 weeks |  |  |
| Sleeping late at night | 0.676 (0.455–1.005) | **0.053** |
| Poor quality of sleep | 0.635 (0.351–1.148) | 0.133 |

OR, odds ratio; 95% CI, 95% confidence interval.

## Discussion

Vaccination using inactivated SARS-CoV-2 is an effective measure to prevent SARS-CoV-2 infection. China, with its large population, provides an opportune study site for assessing vaccination using inactivated SARS-CoV-2 in pregnant women. To our knowledge, this is the first study to investigate the association of sleep status and seroconversion after vaccination using inactivated SARS-CoV-2 in pregnant women. The present study found sleep status was found to be associated with serum level of IgG antibodies.

To explore the impact of sleep status on antibody levels in pregnant women after vaccination against SARS-CoV-2 is a rational approach. Physical discomfort, anxiety, or hormonal changes caused by pregnancy can lead to sleeping problems in pregnant women^[17].^ Studies have shown that disruption of sleep patterns leads to endocrine disorders and changes in the balance of the immune system, including changes in the number of immune cells and cytokine levels in peripheral blood. This phenomenon results in reduced immune defense against, and increased susceptibility to, pathogens^[6]^, which increases the risk of SARS-CoV-2 infection in pregnant women. Which was similar to our findings that pregnant women with poor quality of sleep or SLaN had a lower serum level of IgG antibodies.

The level of the immune response after vaccination is influenced by different factors, including vaccine dose and age, which may have a significant impact on the efficacy and duration of vaccine protection. However, few studies have explored the effect of sleep on the immune response after vaccination with inactivated SARS-CoV-2. In the present study, poor sleep quality was found to be a risk factor for the serum level of IgG antibodies, a finding consistent with that of other vaccine-related studies. In a large-sample cohort study, all participants received two doses of the mRNA SARS-CoV-2 vaccine, but pregnant women were excluded. That study found that poor quality of sleep was associated negatively with antibody levels^[5]^.

Observations on the immune response to vaccines and the relationship with sleep arose originally from animal studies. Brown and colleagues found that clearance of the influenza virus was slowed in sleep-deprived mice compared with that in mice with a normal sleeping pattern^[7]^. Clinical research on sleep and the immune response to vaccination has focused on vaccines against influenza, hepatitis, and *Salmonella* Typhi. A study of a healthy population aged 40–60 years who received three doses of hepatitis-B vaccine revealed that a short duration of sleep was strongly associated with the likelihood of a reduced serum level of antibodies to the hepatitis-B vaccine. Also, this negative effect of sleep disturbance persisted even if triple doses of the vaccine were given, as well as a 6-month adjuvant dose^[8]^. A study of the immune effect of influenza vaccination in college students revealed that poor quality of sleep may be a risk factor for reduced immunity to an influenza virus^[13]^. Appropriately prolonged nighttime sleep after vaccination promotes host immune responses^[14,15]^, and serum-specific antibody concentrations have been found to be significantly lower in sleep-deprived men 5 days after vaccination with the influenza A (HIN1) virus compared with those in a group with a normal sleeping pattern^[9]^. Similarly, people who had a regular sleeping pattern after Hepatitis A virus vaccination had higher antibody titers after 4 weeks^[10]^. One study found that 10 days after vaccination with a seasonal influenza virus, individuals who had 4 consecutive nights of sleep restricted to 4 h had IgG antibody titers less than half that of individuals without this sleep deficit^[16]^.

The body has a “biological clock” to maintain the rhythm of sleep, and SLaN can disrupt this rhythm and affect sleep quality. A recent study showed that frequent SLaN greatly disrupted the ratio of immune cells, and that this alteration contributed to a reduction in the function of immune cells^[18]^. In the present study, SLaN was found to be an independent influence on the serum level of IgG antibodies in pregnant women, with those who SLaN habitually having a lower serum level of IgG antibodies. In addition, the effect of poor sleep quality and SLaN on the serum level of IgG antibodies was found mainly in women aged <35 years. This finding may have been due to the fact that older pregnant women pay more attention to the regularity of their work-and-rest schedule because their body functions decline with age. Napping can reduce drowsiness and may have some benefits for mood stability and cognitive function. An association between napping and antibody levels has been documented, but this finding did not hold after adjustment for confounding factors ^[5]^, which is similar to our findings.

We also identified significant differences between age, time since vaccination,and the serum level of IgG antibodies in pregnant women who received two doses of inactivated SARS-CoV-2 vaccine. Age was an important factor maybe to influencing the serum level of IgG antibodies, and that positivity to IgG antibodies was lower in older pregnant women. A phase-II clinical trial of the CoronaVac vaccine involving 600 healthy adults (18-59 years) revealed that levels of neutralizing antibodies decreased with increasing age^[11]^. Similarly, another study found that the level of the immune response in pregnant women who received an mRNA vaccine was correlated negatively with maternal age^[12]^. Antibody levels in people decrease over time after completing vaccination. One study revealed that antibody levels were correlated negatively with time, and approached the threshold of seropositivity 6 months after receipt of two doses of a vaccine ^[11]^.

This was the first study to analyze the effect of sleep status on the immune response of pregnant women after vaccination with inactivated SARS-CoV-2 based on real-world data in three hospitals in China. However, we declare some limitation in present study. This is a real-world study, which cannot be designed as precisely as a clinical trial. Therefore, data quality may vary widely. Future prospective cohorts with large samples may be needed to further validate the relationship between sleep and the effects of immunization using vaccines.

## Conclusions

Sleep quality and SLaN can affect the serum level of IgG antibodies in pregnant women after vaccination with inactivated SARS-CoV-2.

## Data Availability

All relevant data are within the manuscript and its Supporting Information files.

## Acknowledgments

The authors would like to acknowledge and thank for all people’s contributions during the early stages of the work on this manuscript.

## Notes

### Competing Interest Statement

The authors have declared no competing interest.

### Funding Statement

This study was funded by Exposure to SARS-CoV-2 vaccine before or during pregnancy and adverse pregnancy outcomes: a cohort study(Grant No.41-43241529).

### Author Declarations

The study protocol was approved (IRB 2021-01) by the Medical Ethics Review Board of the School of Public Health within Guangdong Pharmaceutical University (Guangzhou, China). The study protocol complied with the Declaration of Helsinki 1964 and its later amendments.

